# A Propensity-Matched Nested Case-Control Study of Acute Coronary Syndrome Patients Genotyped for CYP2C19

**DOI:** 10.1101/2021.06.30.21259298

**Authors:** Kate Kilpatrick, Nick James, Kevin Smith, John Mackay, Phillip Shepherd, Luke Boyle, Quentin Thurier, Zina Ayar, Patrick Gladding

## Abstract

**Introduction:** Ticagrelor is widely considered superior to clopidogrel however a pharmacogenetic substudy of PLATO indicated that the majority of this difference is due to genetic nonresponders to clopidogrel. We evaluated patient outcomes following genotyping for CYP2C19 in a propensity matched acute coronary syndrome cohort treated with either clopidogrel, ticagrelor or aspirin monotherapy.

**Methods:** ICD10 coding identified 6,985 acute coronary syndrome patients at Waitematā District Health Board over a five year period (2012-2016). Ticagrelor was subsidised by The Pharmaceutical Management Agency of New Zealand in July 2013. Patients were genotyped for CYP2C19 *2, *3 and *17 alleles using the Nanosphere Verigene analyser and treatment was tailored accordingly. Logistic regression and nearest neighbour propensity matching was employed in a 1:3 fashion with each treatment group to balance patient characteristics.

**Results:** A total of 146 patients were genotyped and compared with 438 matched patients taking either clopidogrel, ticagrelor or aspirin monotherapy. Post July 2013 clopidogrel was prescribed more often in responders than in those without genotype information (68 vs 39%, χ2 9, 95% CI 4 to 34, p=0.003). Conversely, ticagrelor was used more frequently in clopidogrel nonresponders. Mortality with personalised treatment was equivalent to ticagrelor (HR 0.8, 95% CI 0.3 to 1.8) but higher in those treated with clopidogrel (HR 2.3, 95 % CI 1 to 5.3). Readmissions with ACS were higher in nonresponders treated with clopidogrel versus those treated with genotype appropriate dual antiplatelet therapy (HR 3.9, 95% CI 0.8 to 18, p =0.03).

**Conclusion:** Personalised antiplatelet management was equivalent to ticagrelor with respect to all-cause mortality and ACS readmissions. It also led to more appropriate use of both clopidogrel and ticagrelor and potential cost savings.

## Introduction

Dual anti-platelet therapy (DAPT) with a platelet P2Y12-receptor inhibitor (ticagrelor, clopidogrel, or prasugrel) and aspirin is the recommended treatment for preventing recurrent ischaemic events and stent thrombosis in acute coronary syndromes (ACS).^1,2^ Clopidogrel is a prodrug that is absorbed in the intestine and activated in the liver. Activation involves two oxidative steps which requires contribution from several different isoenzymes to be converted to its active form. ^2^ The CYP2C19 enzyme has the greatest effect on this process and is an important determinant of pharmacodynamic response to clopidogrel. Individuals carrying loss of function alleles (e.g. *CYP2C19*2, *3*), are considered ‘non responders’ and have a higher risk of ischaemic events and stent thrombosis. This risk is highest in homozygotes for either *2 or *3 variants (poor metabolisers (PMs)).^2^ Conversely, individuals with gain of function alleles (*CYP2C19*17*) are associated with higher rates of bleeding.^3^ Other genes such as ABCB1 which encodes a P-glycoprotein efflux transporter also alters clopidogrel effect through regulating clopidogrel absorption.^4^ Despite a Food and Drug Administration boxed warning, the American College of Cardiology/American Heart Association (ACC/AHA), the European Society of Cardiology (ESC) guidelines and the Clinical Pharmacogenetics Implementation Consortium (CPIC) recommendations on pharmacogenetic testing has not become part of routine care. ^2,5,6^

The PLATO study showed that ticagrelor reduced mortality among patients with ACS, with a risk benefit ratio for bleeding which was considered clinically acceptable.^7^ Although the use of ticagrelor obviates the need for pharmacogenetic testing, compared to clopidogrel ticagrelor is associated with a higher risk of bleeding, increased cost, twice daily administration and adverse effects such as dyspnoea and bradycardia.^8-10^ Although ticagrelor is often used in DAPT in patients with ACS, there remains a role for clopidogrel in patients with adverse reactions to ticagrelor, an increased risk of bleeding, concomitant anticoagulant use, and where affordability is an issue. Switching therapy or de-escalating therapy to clopidogrel or the rational use of prasugrel has been shown to be safe and effective in the PRAGUE-18 study.^11^ In addition a substudy of TROPICAL-ACS showed that genotype information could be used for rationalising DAPT to once daily therapy.^12^

Although randomised clinical trial (RCT) data to support CYP2C19 genotyping has been lacking, recent real-world and pseudo experimental design studies of CYP2C19 pharmacogenetics used in practice have shown promising results.^13-16^ The PHARMCLO study, an RCT, demonstrated that prospective pharmacogenetic testing may reduce ischaemic and bleeding events compared to standard of care, where clopidogrel is the dominant agent of choice.^17^ Here we report a propensity-matched cohort study of a pharmacogenetics program for CYP2C19, evaluating mortality and ACS readmissions. This was performed across a period where ticagrelor was initially not available in patients with ACS.

## Methods

### Study Design & Ethics

This study is a propensity matched retrospective cohort study. Ethics approval was obtained from Awhina Research & Knowledge at Waitematā District Health Board. Registration #RM13622. Support was also obtained from the Waitematā and Auckland District Health Boards Māori Research Committee in December 2016.

### Patients

Patients with ACS admitted to North Shore Hospital (Waitematā District Health Board) from January 1^st^ 2012 until 30^th^ December 2016 were considered eligible for inclusion in the study. A SQL search criteria for ACS using ICD-10 codes was performed (Table 1, Appendix). The CYP2C19 pharmacogenetic program was established in 2012, however ACS patients were not genotyped until February 2013. There were no specific exclusion criteria other than established contraindications and precautions for the use of DAPT (see Treatment). Ethnicity was self-identified at the point of admission.

**Table 1:** Baseline Characteristics.

|  | Ticagrelor<br>n=438 | Clopidogrel<br>n=438 | Aspirin<br>n=438 | Genotyped<br>n=146 |
| --- | --- | --- | --- | --- |
| Age (mean ((SD)) | 64 (12) | 64 (13) | 65 (12) | 64 (13) |
| Weight (kg) | 85 (18) | 87 (22) | 86 (23) | 82 (21) |
| Male (%) | 279 (64) | 298 (68) | 304 (69) | 98 (67) |
| Hypertension (%) | 272 (62) | 269 (61) | 270 (62) | 89 (61) |
| Diabetes Mellitus (%) | 93 (21) | 94 (21) | 109 (25) | 32 (22) |
| Dyslipidaemia (%) | 39 (9) | 56 (13) | 52 (12) | 18 (12) |
| Smoking (%) | 253 (58) | 260 (59) | 255 (58) | 84 (58) |
| Prior atherosclerosis (%) | 275 (63) | 255 (58) | 219 (50) | 88 (60) |
| Renal impairment (%) | 29 (7) | 19 (4) | 28 (6) | 9 (6) |
| PCI (%) | 115 (35) | 123 (35) | 21 (8)* | 44 (36) |

PCI data was incomplete in approximately 20% of cases \*p<0.05

### Treatment

Patients were treated according to physician discretion and medication availability. Ticagrelor was funded by The Pharmaceutical Management Agency of New Zealand on July 1^st^ 2013. Prasugrel was funded from 1^st^ April 2012 for clopidogrel-allergic patients who have undergone percutaneous coronary intervention (PCI). Local hospital guidelines for the use of DAPT recommend that patients with ST elevation and non-ST elevation myocardial infarction (MI) are preferably treated with ticagrelor. Whereas patients with type II MI, those with low ischaemic risk (troponin <40ng/L, no ischaemic changes on ECG with diagnosed unstable angina) or high bleeding risk (anaemia, recent bleeding, high CRUSADE bleeding score,^18^ age >80 years or weight <60kg) are treated with clopidogrel. Ticagrelor is contraindicated in patients treated with oral anticoagulants (warfarin or dabigatran), recent thrombolysis (<24 hr) or previous intra-cranial bleed. It is not recommended in those with severe heart failure, severe conduction disease without a permanent pacemaker or severe asthma. Genotyped patients were treated according to CPIC guidelines with the addition of clinical discretion. An antiplatelet algorithm (PREDICT) including age, renal dysfunction, heart failure and genotype information was available in an electronic form to guide personalised care for each patient.^5,19^

### Genotyping

Single nucleotide polymorphism (SNP) genotyping was performed using the Nanosphere Verigene Analyser (Luminex, Austin, Texas). *CYP2C19*2* and *\*3* loss of function alleles, and *\*17* gain of function allele were genotyped within a 2 hour time frame, with results reported electronically using an algorithm based on CPIC guidelines.^5^ Patients heterozygous for *\*2* or *\*3* or with *\*2/*17* polymorphisms (Intermediate metabolisers (IMs)) were recommended to receive clopidogrel 225mg once daily, ticagrelor or prasugrel. Patients homozygous for *\*2* or *\*3* (poor metabolisers (PMs)) were recommended to receive ticagrelor or prasugrel when available. Patients homozygous for *17 (ultrametabolisers (UMs)) were given a precaution for bleeding, particularly in the context of concomitant anticoagulant use.

Clopidogrel nonresponders = poor metabolisers (PMs) and intermediate metabolisers (IMs). PMs = *\*2/*2, *2/*3, *3/*3*; IMs = *\*2/wild type (wt), *3/wt, *17/*2*. Clopidogrel responders = extensive metabolisers (EMs). EMs = *wt/wt*. Ultra metabolisers (UMs) = *\*17/*17*.

### Outcomes

Genotype frequency across varying ethnicity was measured. Prescribing behaviour was monitored using inpatient Pyxis records (Pyxis, San Diego), however clopidogrel dose and discharge medications could not be evaluated. The primary clinical outcome was all-cause mortality over a twelve month period from discharge after myocardial infarction. Cardiovascular mortality could not be identified through hospital electronic clinical records. The secondary outcome was readmission due to ACS over a twelve month period identified by ICD10 codes. Outcomes were analysed on an intention to treat basis including genotypes with no calls (a failed test) for mortality and readmissions. When analysed by specific genotype no calls were excluded.

### Propensity Matching

To estimate the propensity scores, we partitioned the cohort into three groups; 1) a group consisting of all genotyped and all aspirin-only treated patients, 2) a group consisting of all genotyped patients and all clopidogrel patients, and 3) a group consisting of all genotyped patients and all ticagrelor/prasugrel patients. A fitted logistic regression model was then developed on each group in which the outcome was the log-odds of being genotyped. The independent variables (covariates) for each logistic model included age, gender, diabetes, renal impairment, smoking, hypertension and dyslipidaemia.

Matching was then employed within each of the three groups to select control patients (the aspirin, clopidogrel, and ticagrelor treatments) who were similar to patients that were genotyped (the experimental treatment). As the sample size for each control group was considerably larger than the sample size for the experimental treatment (6 – 20 times as many samples in each control treatment), we conducted a 1:3 (1 experimental patient matched to 3 control patients) greedy nearest-neighbour matching scheme within each group, with a fixed caliper width of 0.25. Control patients that were within the fixed caliper width were matched at random with the experimental patients. Control patients selected by the matching algorithm were included within the matched sub-cohort for each of the three groups and patients not selected were discarded. Patients from the three matched control treatments (aspirin, clopidogrel, and ticagrelor/prasugrel) were then merged into a single cohort along with the patients from the experimental treatment.

### Statistics

Unpaired student t tests were used to compare baseline characteristics, ethnic and genotype frequencies. Chi squared testing was used to compare proportions. Kaplan Meier curves and hazard ratios were used to evaluate both mortality and readmission outcomes. Cox-proportional hazard models were used to evaluate clinical factors which influenced outcomes. Statistical analysis was performed using MedCalc 17.7.2.

## Results

### Patients

ICD10 coding identified 6,985 ACS patients at Waitematā District Health Board over a five-year period (2012-2016). A total of 146 patients were genotyped and compared with 438 propensity matched patients taking either clopidogrel, ticagrelor or aspirin monotherapy. Baseline characteristics are shown in Table 1 with no statistically significant differences between groups due to effective propensity matching. Ethnicity mix within the propensity matched cohorts were similar to the genotyped cohort (Table 2).

**Table 2:** Ethnic Mix Between Cohorts.

|  | Ticagrelor<br>n=438 | Clopidogrel<br>n=438 | Aspirin<br>n=438 | Genotyped<br>n=146 |
| --- | --- | --- | --- | --- |
| European (%) | 310 (71) | 316 (72) | 319 (73) | 101 (69) |
| Asian (%) | 42 (10) | 47 (11) | 47 (11) | 16 (11) |
| Pacific Island (%) | 38 (9) | 29 (7) | 28 (6) | 17 (12) |
| Māori (%) | 31 (7) | 32 (7) | 37 (8) | 9 (6) |
| Other (%) | 17 (4) | 14 (3) | 7 (2) | 3 (2) |

Includes patients with no calls.

### Prescribing behaviour

Including patients with no calls, 92 genotyped patients (63%) were treated with clopidogrel, 36 with ticagrelor (25%), 1 with prasugrel (0.7%) and 17 with either aspirin or no treatment was recorded (12%). Eight (73%) of the clopidogrel nonresponders were treated with clopidogrel prior to July 2013 versus 22 (49%) post July 2013 (χ2 2, 95% CI -13 to 50, p =0.16). Post July 2013, 44 (68%) clopidogrel responders versus 1730 (39%) without genotype information were prescribed clopidogrel (χ2 23, 95% CI 16 to 40, p<0.0001). Conversely ticagrelor was used more frequently in clopidogrel nonresponders (16, 36%) versus those without genotype information (886, 20%) (χ2 7, 95% CI 2 to 32, p =0.009), though there was a trend towards greater ticagrelor use across the whole cohort over time (Table 3, Figure 1).

**Table 3:**
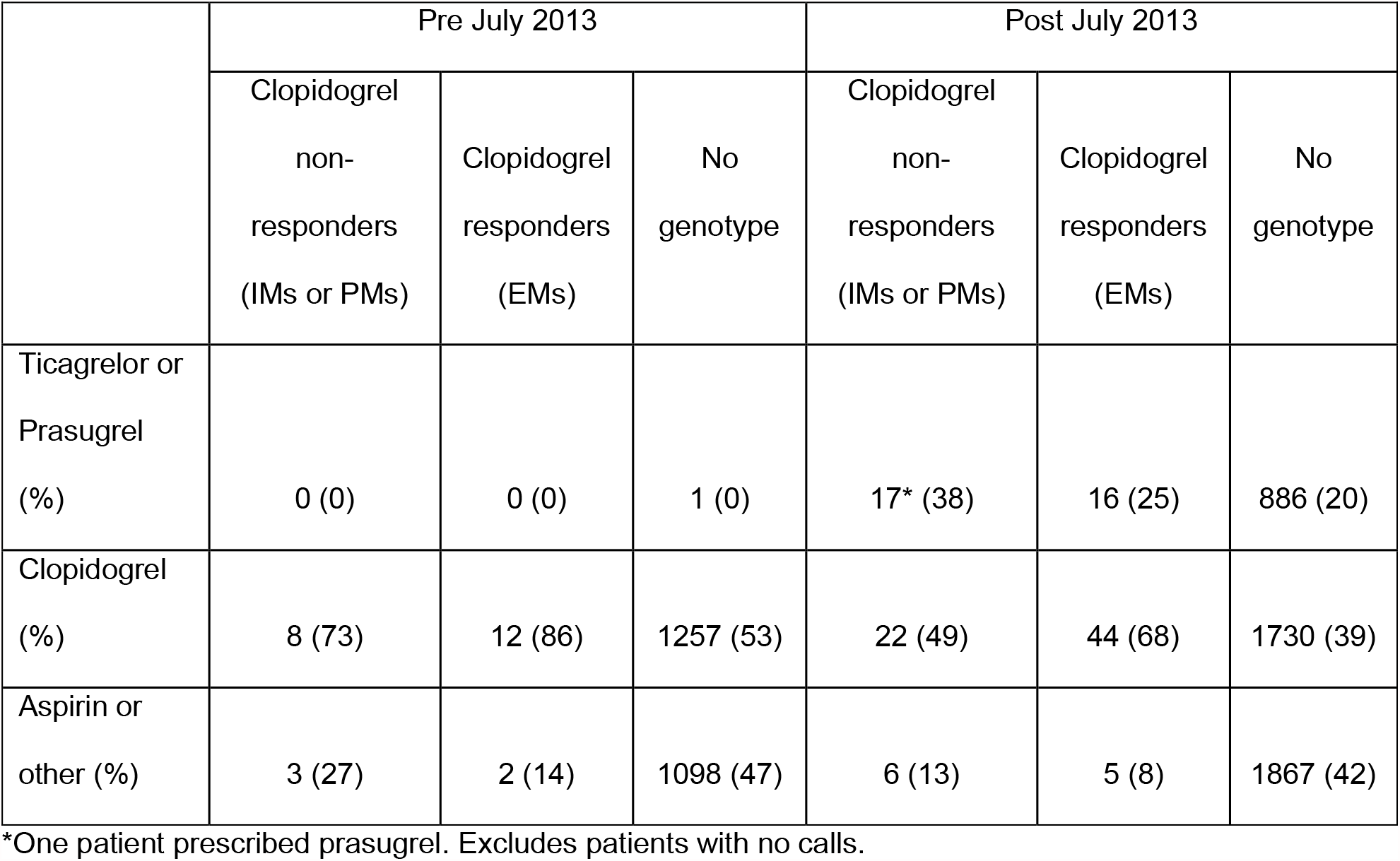
Genotyping Status and Prescribing Behaviour.

|  | Pre July 2013 |  |  | Post July 2013 |  |  |
| --- | --- | --- | --- | --- | --- | --- |
|  | Clopidogrel<br>non-<br>responders<br>(IMs or PMs) | Clopidogrel<br>responders<br>(EMs) | No<br>genotype | Clopidogrel<br>non-<br>responders<br>(IMs or PMs) | Clopidogrel<br>responders<br>(EMs) | No<br>genotype |
| Ticagrelor or<br>Prasugrel<br>(%) | 0 (0) | 0 (0) | 1 (0) | 17* (38) | 16 (25) | 886 (20) |
| Clopidogrel<br>(%) | 8 (73) | 12 (86) | 1257 (53) | 22 (49) | 44 (68) | 1730 (39) |
| Aspirin or<br>other (%) | 3 (27) | 2 (14) | 1098 (47) | 6 (13) | 5 (8) | 1867 (42) |

\*One patient prescribed prasugrel. Excludes patients with no calls.

**Figure 1:**
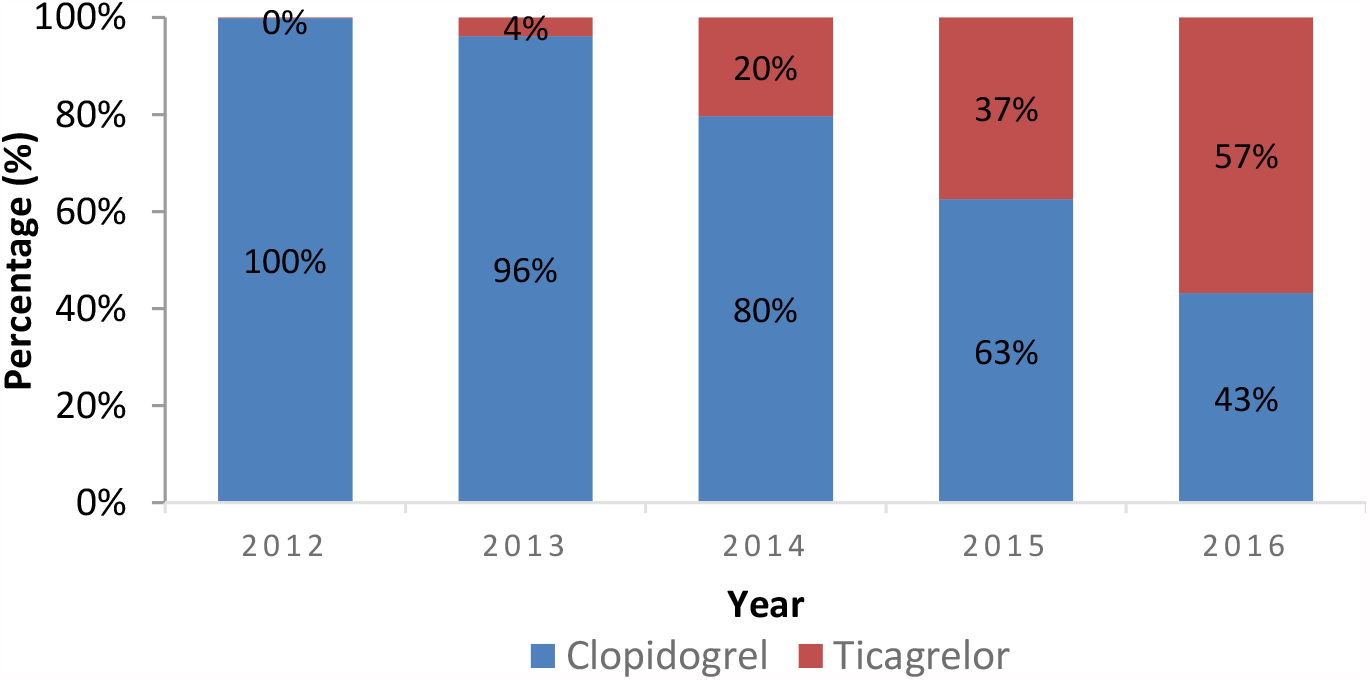
Proportion of Ticagrelor and Clopidogrel In-Patient Prescriptions at Waitematā District Health Board 2012-2016.

### Genotype and Ethnic frequency

The number of patients genotyped were low across the study period (Figure 2). Eleven (8%) of the Nanosphere Verigene Analyser genotype results returned as “no call”. Of the remaining 135 genotyped patients the ethnic distribution of genotype frequencies are shown in Table 4. Pacific Peoples had the highest frequency of clopidogrel nonresponders followed by Asians. Māori had a similar frequency of clopidogrel nonresponders to Europeans.

**Figure 2:**
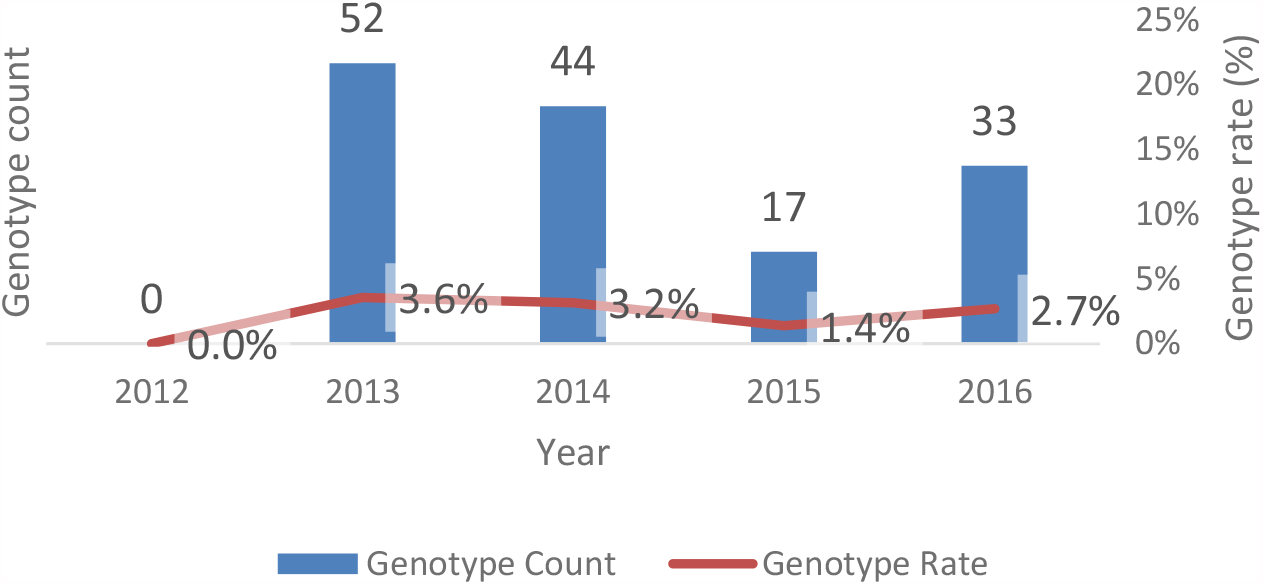
Annual Rates of Genotyping at Waitematā District Health Board 2012-2016.

**Table 4:** Ethnic Distribution of Genotyped Patients.

|  | *Clopidogrel*<br>*non-responders* |  |  | *Clopidogrel*<br>*responders* |  |  |
| --- | --- | --- | --- | --- | --- | --- |
|  | PMs | IMs | PMs or IMs | EMs | UMs | Total |
| European (%) | 3 (3) | 29 (31) | 32 (34) | 59 (63) | 3 (3) | 94 |
| Pacific Island (%) | 0 (0) | 12 (75) | 12 (75)± | 4 (25) | 0 (0) | 16 |
| Asian (%) | 2 (14) | 7 (50) | 9 (64)± | 5 (36) | 0 (0) | 14 |
| Māori (%) | 1 (13) | 2 (25) | 3 (38) | 5 (62) | 0 (0) | 8 |
| Other (%) | 0 (0) | 0 (0) | 0 (0) | 3 (100) | 0 (0) | 3 |
| Total (%) | 6 (4) | 50 (37) | 56 (41) | 76 (57) | 3 (2) | 135 |

Excludes patients with no calls. ± p<0.05.

Clopidogrel nonresponders = poor metabolisers (PMs) and intermediate metabolisers (IMs). PMs = *\*2/*2, *2/*3, *3/*3*; IMs = *\*2/wild type (wt), *3/wt, *17/*2*. Clopidogrel responders = extensive metabolisers (EMs). EMs = *wt/wt*. Ultra-metabolisers (UMs) = *\*17/*17*.

### Mortality

Across the entire propensity matched cohort both Māori and Pacific Peoples had an unadjusted increased risk of mortality compared to Europeans (Māori HR 2.2, 95% CI 0.8 to 5.8; Pacific people HR 1.6, 95% CI 0.6 to 4). Cox-proportional hazard modelling adjusted for baseline clinical characteristics and treatment group showed that age, ethnicity and renal dysfunction accounted for the largest proportion of variability in mortality outcomes (Table 5). However in just the genotyped group only renal dysfunction remained statistically significant. Individual genotypes occurred at too low a frequency to be used in the Cox model.

**Table 5:** Cox-Proportional Hazard Model for Mortality. Adjusted for Baseline Clinical Characteristics Using Propensity Matching.

| Covariate | b | SE | Wald | P | Exp(b) | 95% CI of Exp(b) |
| --- | --- | --- | --- | --- | --- | --- |
| Age | 0.09955 | 0.01334 | 55.6722 | <0.0001 | 1.1047 | 1.0762 to 1.1339 |
| Maori | 1.7308 | 0.3989 | 18.8209 | <0.0001 | 5.6450 | 2.5827 to 12.3383 |
| Pacific Peoples | 0.9397 | 0.4345 | 4.6778 | 0.0306 | 2.5591 | 1.0921 to 5.9966 |
| Renal impairment | 1.4466 | 0.3445 | 17.6378 | <0.0001 | 4.2488 | 2.1630 to 8.3460 |

Mortality was higher in the group prescribed aspirin monotherapy and clopidogrel, compared to either ticagrelor or genotyped patients, though the mortality difference with clopidogrel only reached borderline statistical significance (Figure 3, Table 6). In terms of mortality, personalised care (genotyped) appeared equivalent to ticagrelor.

**Figure 3:**
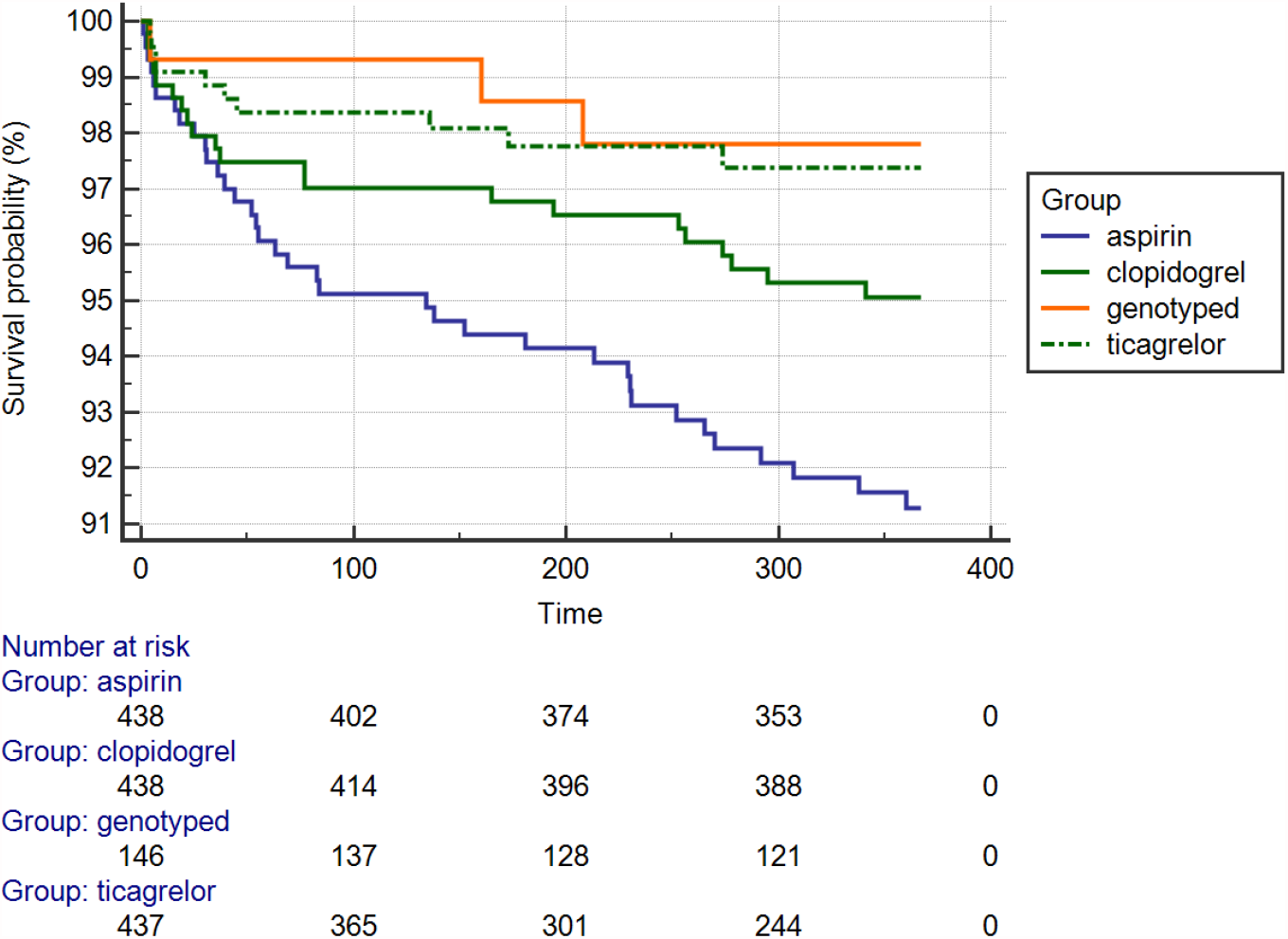
Comparison of Survival Curves for the Aspirin Monotherapy, Clopidogrel, Ticagrelor and Genotyped Groups.

**Table 6:** Comparison of Survival Curves (Logrank Test).

| Factor | aspirin | clopidogrel | genotyped | ticagrelor |
| --- | --- | --- | --- | --- |
| aspirin | - | HR 0.6<br>95 % CI 0.3 to 1 | HR 0.2<br>95 % CI 0.1 to 0.6 | HR 0.3<br>95 % CI 0.2 to 0.6 |
| clopidogrel | 1.8<br>95 % CI 1 to 3.2 | - | HR 0.4<br>95 % CI 0.2 to 1 | HR 0.5<br>95 % CI 0.3 to 1 |
| genotyped | HR 4.1<br>95 % CI 1.8 to 9.5 | HR 2.3<br>95 % CI 1 to 5.3 | - | HR 1.3<br>95 % CI 0.5 to 3 |
| ticagrelor | HR 3.2<br>95 % CI 1.7 to 6 | HR 1.8<br>95 % CI 1 to 3.3 | HR 0.8<br>95 % CI 0.3 to 1.8 | - |

$\chi^2$ 17, Degrees of freedom (DF) 3, P = 0.0007

### Readmissions

Readmissions with ACS appeared higher in the genotyped, aspirin monotherapy and clopidogrel groups compared to ticagrelor however this did not reach statistical significance (HR 3, 95% CI 1.2 to 8, P = 0.08) (Figure 4). Within the genotyped group readmission rates were predominantly driven by clopidogrel nonresponders (IMs or PMs) prescribed clopidogrel, compared to those given treatment appropriate for their genotype and clinical profile (HR 3.9, χ2 5, DF 1, 95% CI 0.8 to 18, p =0.03) (Figure 5). Cox proportional hazard modelling showed that the major covariate of importance was renal dysfunction in predicting readmissions (Table 7). Six patients were identified as PMs i.e. *\*2/*2*, or *\*3/*3* and four of these patients (67%) received either ticagrelor or prasugrel at their index admission. The two PMs treated with clopidogrel, admitted prior to July 2013, had early readmissions within two weeks of their index event. One had a prior history of multiple stent reocclusions whilst on clopidogrel and eventually underwent cardiac transplantation for ischaemic cardiomyopathy. After representation both patients were placed on genotype appropriate DAPT.

**Figure 4:**
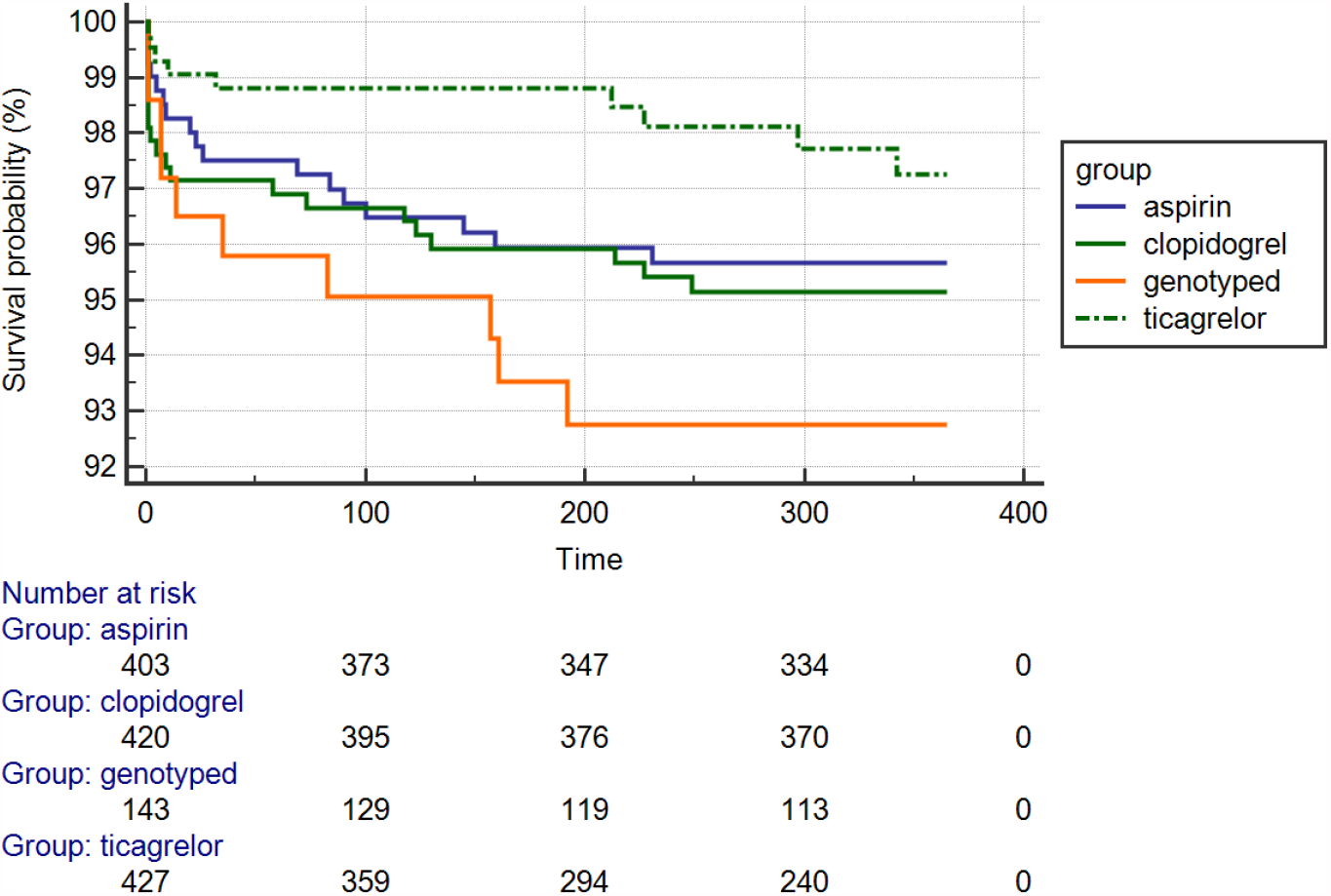
Comparison of Readmission with ACS in the Aspirin Monotherapy, Clopidogrel, Ticagrelor and Genotyped Groups.

**Figure 5:**
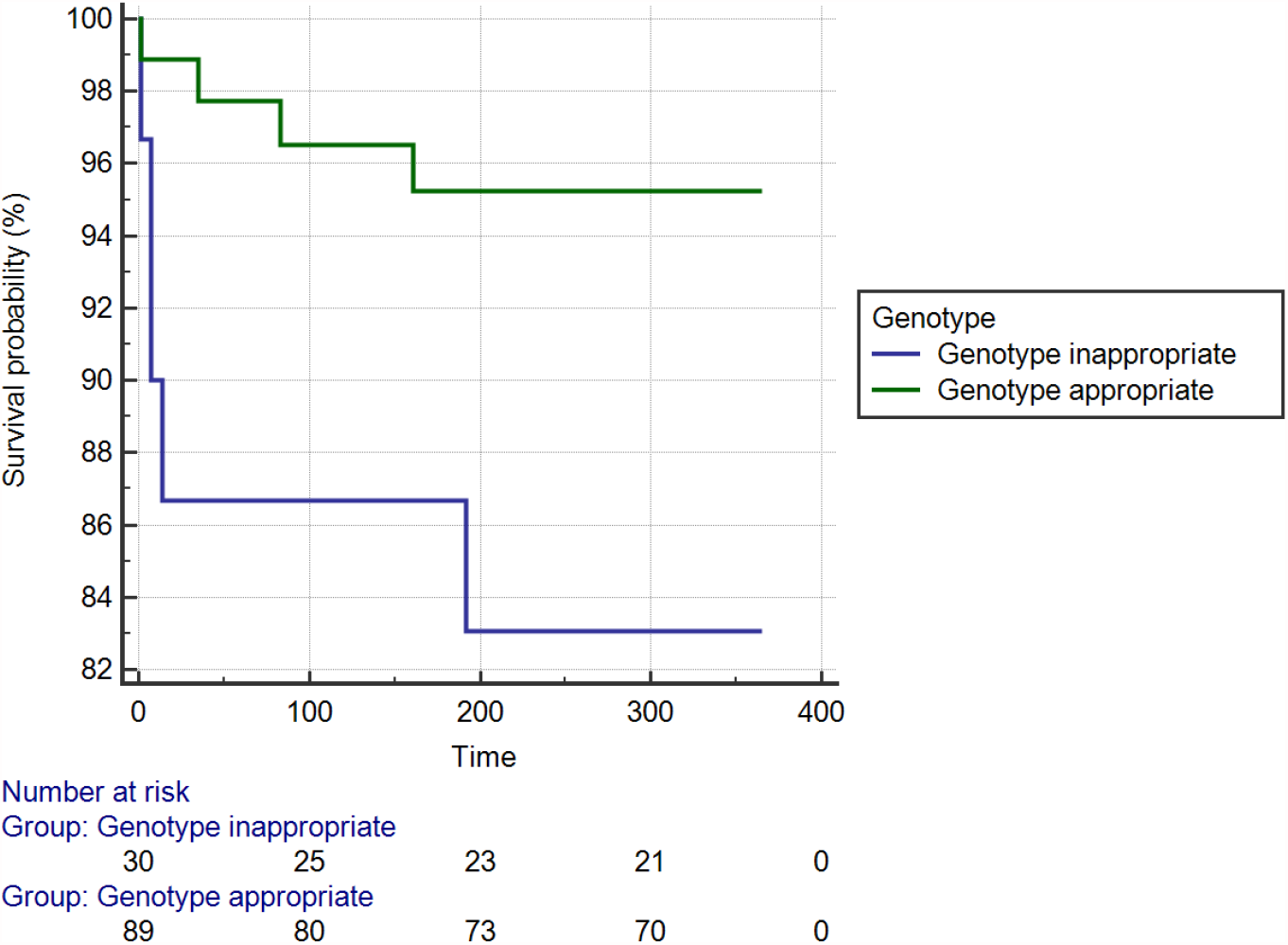
Comparison of Readmissions with ACS in the Genotype Inappropriate Group and the Genotyped Appropriate Treatment Groups.

**Table 7:** Cox Proportional Hazard Model for ACS Readmissions. Adjusted for baseline clinical characteristics using propensity matching.

| Covariate | b | SE | Wald | P | Exp(b) | 95% CI of Exp(b) |
| --- | --- | --- | --- | --- | --- | --- |
| Renal_impairment__AKI_CKD_=1 | 2.8073 | 1.4143 | 3.9399 | 0.0472 | 16.5659 | 1.0359 to 264.9222 |

## Discussion

In this propensity matched observational cohort study we have shown that personalised treatment can be effectively and safely delivered at point of care. Although the numbers of patients genotyped and outcome rates were low, personalised treatment appeared to be equivalent to treatment with ticagrelor with respect to mortality. Superiority of personalised care to clopidogrel was borderline significant for mortality outcomes. Readmissions rates with ACS occurred at a low frequency and were not statistically significant between treatment groups, though there was a trend to higher readmission rates in those that were genotyped. This was driven largely by genetic nonresponders who were treated with clopidogrel. It is worth noting that even when genotype information and ticagrelor was available, 49% of nonresponders are still prescribed clopidogrel. Unfortunately due to the limited information available it was not possible to identify which of these patients received the recommended 225mg clopidogrel dose, so it is unknown whether these patients received treatment appropriate to CPIC guidelines. However the patients with the highest risk of stent thrombosis and reinfarction are PMs who are not able to metabolise clopidogrel 75mg or 225mg. All PMs genotyped after July 2013 received genotype appropriate therapy. We have previously shown a significantly higher rate of stent thrombosis events in patients who are homozygous for either *CYP2C19*2* or *\*3* genotypes.^20^ Although this study was an observational design, with the risk of confounding and bias, we evaluated the real world practice of clinical pharmacogenetics and demonstrated results consistent with previous reports.

The frequency of clopidogrel CYP2C19 non-responder variants is known to be higher in some ethnic groups such as Asians, Pacific People, Māori and African Americans.^21-23^ The ethnicity of those genotyped in this study matched the whole cohort showing that ethnicity targeted genetic testing did not occur. In this cohort the frequency of non-responder variants was higher in Asian and Pacific People but not Māori, probably due to the small sample of Māori tested. Although ethnicity can be used as a partial proxy for genotype, Cox hazard modelling identified only Māori and Pacific ethnicity, but not Asian as independent risk factors for mortality. Unmeasured influences such as socioeconomic factors and deprivation may have accounted for these differences.

Evaluation of the CYP2C19 genotype and its influence on clopidogrel efficacy has now spanned pharmacokinetic, pharmacodynamic, clinical outcome, meta-analysis studies, propensity matched and more recently randomised controlled trials.^24^ Despite this level of evidence and the recommendations in clinical guidelines, genotyping has still not become part of standard of care. In part this is due to some inconsistencies in the literature, often a result of meta-analyses that included inappropriate patient subgroups e.g. patients with atrial fibrillation treated with clopidogrel.^25^ There are still multiple barriers to incorporating pharmacogenetics into standard of care. These include clinician education, availability, cost and further randomised evidence. In our study, despite local evidence supporting the use of genotyping and its availability, testing rates were extremely low. Mostly this occurred due to clinician’s not ordering the test, possibly due to lack of awareness of its utility and distraction with other patient management issues. For these reasons we believe that pharmacist driven implementation may be a better policy initiative than expecting clinicians to remember to order the test. In some centres pre-emptive genotyping has been performed to obviate the need of remembering to order the test. As a result, these centres have amassed data that has provided support for CYP2C19 genotyping, through the monitoring of patient outcomes linked to pharmacogenetic driven decision making.^15^ This provides an excellent example of a learning healthcare system and a prototype for the wider implementation of genomic medicine through pragmatic clinical trials.

Although the ACC/AHA guidelines for ACS recommend the use of genotyping in patients who have suffered reinfarction after ACS,^2,26^ it would seem more practical that pharmacogenetic information be used to prevent secondary events. Although the number of poor metabolisers in this study were low it is worth noting that the two CYP2C19 non-responder homozygotes treated with clopidogrel were readmitted with ACS within two weeks from discharge. Furthermore, one of these patients had a long history of serial stent occlusions which resulted in a severe ischaemic cardiomyopathy requiring cardiac transplantation. It is plausible that this transplantation could have been avoided with pre-emptive pharmacogenetic testing.

Several cost-effectiveness analyses have been performed for CYP2C19 genetic testing and antiplatelet use. Although some studies have shown mixed results, in general the majority of these analyses have shown that genetic testing is cost-effective when compared to one-size-fits all use of ticagrelor, which is substantially more expensive than generic clopidogrel.^13,27-34^ Furthermore few or none of these analyses have modelled the effect of non-adherence, poor persistence or the cost of investigations for dyspnoea caused by ticagrelor. The limitation of these cost-effectiveness analyses is that they have been unable to incorporate the rapidly reducing cost in obtaining genomic information. Genomic information has become exponentially less costly over time, especially when delivered at scale. The Verigene analyser used in this study provided a result in less than two hours for USD$100, however we have subsequently validated this against an iPLEX assay, using the Sequenom (Agena Bioscience, San Diego) costing $60, and more recently RT-PCR using a BDMax costing $30/test. Similarly genome wide SNP microarrays, such as the UK Biobank Axiom array, cost only USD$50 and provide a wealth of additional polygenic risk data for cardiovascular events, atrial fibrillation, lipid profile and non-cardiovascular disease such as type II diabetes and cancer. ^35^ Such arrays are also able to generate SNP data for other pharmacogenes involved with clopidogrel, prasugrel and ticagrelor metabolism e.g. ABCB1, PON1, CES and other CYP450 genes.^36-40^

It appears that genetic clopidogrel non-responders with the greatest risk are those with ACS who have undergone PCI and less so those undergoing elective PCI.^24^ Clinical concomitant factors which further increase the risk of stent thrombosis include diabetes, renal dysfunction, left ventricular systolic dysfunction and age. Algorithms such as the PREDICT tool used in this study integrate clinical factors such as these with genetic information to make the most appropriate treatment decision for the individual patient.^19^ Other pharmacogenetic algorithms exist which include stent characteristics and other factors, however their clinical implementation is limited by complexity and availability.^19,41-44^ Clinical tools such as the DAPT score^45^ attempt to balance the risk of reinfarction against bleeding to determine the duration of DAPT however it could be envisaged that a single decision support tool, integrating genetic information, used at the point of discharge could advise the best course of treatment for each patient. Ideally this would involve once daily administration to improve adherence, balance ischaemic against bleeding risk and determine duration of treatment. It seems intuitive that maintenance DAPT should be less intensive than upfront DAPT at the time of admission with ACS, however robust evidence for this assertion is currently lacking. The PRAGUE-18 study^11^ and TROPICAL-ACS genetic substudy^12^ support the concept of de-escalation of DAPT however this hypothesis will need to be tested in a RCT. As genetic information becomes more available it would also seem probable that non-pharmacogenetic risk factors for reinfarction would also be incorporated into these decision support tools. Pragmatic randomised registries or clinical trials, with feedback loops embedded in clinical care would be the ideal method for developing real-time decision support algorithms. With current electronic health records and machine learning tools this prospect seems entirely feasible but will require commitment from clinicians and investment from payers.

The main limitation of this study is that it was observational and therefore prone to external bias. The method of data retrieval using electronic ICD10 coding is not always reliable and medication doses at discharge were not available. It was also not clear why a substantial proportion of patients received either aspirin monotherapy or had no recorded second antiplatelet agent. Whilst it is probable that a number of these would either have had bleeding contraindications or been on anticoagulants the proportion of patients with unidentified DAPT was greater than we would expect. The numbers of patients genotyped in this study was low requiring unbalanced propensity matching to compensate. A greater number of patients genotyped would be required to adequately assess mortality outcomes to compare personalised care with one-size-fits all ticagrelor.

Despite this, we have demonstrated that the clinical implementation of pharmacogenetic testing for CYP2C19 clopidogrel non-responder variants is feasible, with results delivered in a timely fashion to impact on clinical care. Whilst not all genetic information altered treatment decisions, patients who had treatment appropriate to genotype and clinical factors were less likely to suffer reinfarction in a twelve month period and overall had similar mortality to those treated with one-size-fits all ticagrelor in the absence of genetic information.

## Data Availability

The data that support the findings of this study are available on request from the corresponding author upon approval of data sharing committees of the respective institutions.

## Acknowledgements

This research was supported by the University of Auckland and Precision Driven Health.

## References

1. Roffi M, Patrono C, Collet JP, et al. 2015 ESC Guidelines for the management of acute coronary syndromes in patients presenting without persistent ST-segment elevation: Task Force for the Management of Acute Coronary Syndromes in Patients Presenting without Persistent ST-Segment Elevation of the European Society of Cardiology (ESC). Eur Heart J. 2016;37(3):267–315.

2. Amsterdam EA, Wenger NK, Brindis RG, et al. 2014 AHA/ACC Guideline for the Management of Patients with Non-ST-Elevation Acute Coronary Syndromes: a report of the American College of Cardiology/American Heart Association Task Force on Practice Guidelines. J Am Coll Cardiol. 2014;64(24):e139–e228.

3. Sibbing D, Koch W, Gebhard D, et al. Cytochrome 2C19*17 allelic variant, platelet aggregation, bleeding events, and stent thrombosis in clopidogrel-treated patients with coronary stent placement. Circulation. 2010;121(4):512–8.

4. Mega JL, Close SL, Wiviott SD, et al. Genetic variants in ABCB1 and CYP2C19 and cardiovascular outcomes after treatment with clopidogrel and prasugrel in the TRITON-TIMI 38 trial: a pharmacogenetic analysis. Lancet. 2010;376(9749):1312–9.

5. Scott SA, Sangkuhl K, Gardner EE, et al. Clinical Pharmacogenetics Implementation Consortium guidelines for cytochrome P450-2C19 (CYP2C19) genotype and clopidogrel therapy. Clin Pharmacol Ther. 2011;90(2):328–32.

6. Roffi M, Patrono C, Collet JP, et al. 2015 ESC Guidelines for the management of acute coronary syndromes in patients presenting without persistent ST-segment elevation: Task Force for the Management of Acute Coronary Syndromes in Patients Presenting without Persistent ST-Segment Elevation of the European Society of Cardiology (ESC). Eur Heart J. 2016;37(3):267–315.

7. Wallentin L, Becker RC, Budaj A, et al. Ticagrelor versus clopidogrel in patients with acute coronary syndromes. N Engl J Med. 2009;361(11):1045–57.

8. Becker RC, Bassand JP, Budaj A, et al. Bleeding complications with the P2Y12 receptor antagonists clopidogrel and ticagrelor in the PLATelet inhibition and patient Outcomes (PLATO) trial. Eur Hear J. 2011;32(23):2933–44.

9. Storey RF, Becker RC, Harrington RA, et al. Characterization of dyspnoea in PLATO study patients treated with ticagrelor or clopidogrel and its association with clinical outcomes. Eur Heart J. 2011;32(23):2945–53.

10. Cattaneo M, Schulz R, Nylander S. Adenosine-mediated effects of ticagrelor: evidence and potential clinical relevance. J Am Coll Cardiol. 2014;63(23):2503–9.

11. Motovska Z, Hlinomaz O, Kala P, et al. 1-Year Outcomes of Patients Undergoing Primary Angioplasty for Myocardial Infarction Treated With Prasugrel Versus Ticagrelor. J Am Coll Cardiol. 2018;71(4):371–81.

12. Gross L, Trenk D, Jacobshagen C, et al. Genotype-Phenotype Association and Impact on Outcomes following Guided De-Escalation of Anti-Platelet Treatment in Acute Coronary Syndrome Patients: The TROPICAL-ACS Genotyping Substudy. Thromb Haemost. 2018.

13. Deiman BA, Tonino PA, Kouhestani K, et al. Reduced number of cardiovascular events and increased cost-effectiveness by genotype-guided antiplatelet therapy in patients undergoing percutaneous coronary interventions in the Netherlands. Neth Heart J. 2016;24(10):589–99.

14. Sanchez-Ramos J, Davila-Fajardo CL, Toledo Frias P, et al. Results of genotype-guided antiplatelet therapy in patients who undergone percutaneous coronary intervention with stent. Int J Cardiol. 2016;225:289–95.

15. Cavallari LH, Lee CR, Beitelshees AL, et al. Multisite Investigation of Outcomes With Implementation of CYP2C19 Genotype-Guided Antiplatelet Therapy After Percutaneous Coronary Intervention. JACC Cardiovasc Interv. 2018;11(2):181–91.

16. Lee CR, Sriramoju VB, Cervantes A, et al. Clinical Outcomes and Sustainability of Using CYP2C19 Genotype-Guided Antiplatelet Therapy After Percutaneous Coronary Intervention. Circ Genom Precis Med. 2018;11(4):e002069.

17. Notarangelo FM, Maglietta G, Bevilacqua P, et al. Pharmacogenomic Approach to Selecting Antiplatelet Therapy in Patients With Acute Coronary Syndromes: The PHARMCLO Trial. J Am Coll Cardiol. 2018;71(17):1869–77.

18. Subherwal S, Bach RG, Chen AY, et al. Baseline risk of major bleeding in non-ST-segment-elevation myocardial infarction: the CRUSADE (Can Rapid risk stratification of Unstable angina patients Suppress ADverse outcomes with Early implementation of the ACC/AHA Guidelines) Bleeding Score. Circulation. 2009;119(14):1873–82.

19. Geisler T, Schaeffeler E, Dippon J, et al. CYP2C19 and nongenetic factors predict poor responsiveness to clopidogrel loading dose after coronary stent implantation. Pharmacogenomics. 2008;9(9):1251–9.

20. Sathananthan J, El-Jack S, Khan A, et al. TCT-503 Prevalence of CYP2C19 Variants and Associated Stent Thrombosis in Patients Undergoing Percutaneous Coronary Intervention. J Am Coll Cardiol. 2014;64(11 Supplement):B148.

21. Lea RA, Roberts RL, Green MR, et al. Allele frequency differences of cytochrome P450 polymorphisms in a sample of New Zealand Maori. N Z Med J. 2008;121(1272):33–7.

22. Martis S, Peter I, Hulot JS, et al. Multi-ethnic distribution of clinically relevant CYP2C genotypes and haplotypes. Pharmacogenomics J. 2013;13(4):369–77.

23. Helsby NA. CYP2C19 and CYP2D6 genotypes in Pacific peoples. Br J Clin Pharmacol. 2016;82(5):1303–7.

24. Crews KR, Gaedigk A, Dunnenberger HM, et al. Clinical Pharmacogenetics Implementation Consortium guidelines for cytochrome P450 2D6 genotype and codeine therapy: 2014 update. Clin Pharmacol Ther. 2014;95(4):376–82.

25. Holmes MV, Perel P, Shah T, et al. Cyp2c19 genotype, clopidogrel metabolism, platelet function, and cardiovascular events: A systematic review and meta-analysis. JAMA. 2011;306(24):2704–14.

26. Holmes DR, Dehmer GJ, Kaul S, et al. ACCF/AHA clopidogrel clinical alert: approaches to the FDA “boxed warning”: a report of the American College of Cardiology Foundation Task Force on clinical expert consensus documents and the American Heart Association endorsed by the Society for Cardiovascular Angiography and Interventions and the Society of Thoracic Surgeons. J Am Coll Cardiol. 2010;56(4):321–41.

27. Kazi DS, Garber AM, Shah RU, et al. Cost-effectiveness of genotype-guided and dual antiplatelet therapies in acute coronary syndrome. Ann Intern Med. 2014;160(4):221–32.

28. Panattoni L, Brown PM, Te Ao B, et al. The cost effectiveness of genetic testing for CYP2C19 variants to guide thienopyridine treatment in patients with acute coronary syndromes: a New Zealand evaluation. Pharmacoeconomics. 2012;30(11):1067–84.

29. Reese ES, Daniel Mullins C, Beitelshees AL, et al. Cost-effectiveness of cytochrome P450 2C19 genotype screening for selection of antiplatelet therapy with clopidogrel or prasugrel. Pharmacotherapy. 2012;32(4):323–32.

30. Wang Y, Yan BP, Liew D, et al. Cost-effectiveness of cytochrome P450 2C19 *2 genotype-guided selection of clopidogrel or ticagrelor in Chinese patients with acute coronary syndrome. Pharmacogenomics J. 2017.

31. Patel V, Lin FJ, Ojo O, et al. Cost-utility analysis of genotype-guided antiplatelet therapy in patients with moderate-to-high risk acute coronary syndrome and planned percutaneous coronary intervention. J Pharm Pract. 2014;12(3):438.

32. Sorich MJ, Horowitz JD, Sorich W, et al. Cost-effectiveness of using CYP2C19 genotype to guide selection of clopidogrel or ticagrelor in Australia. Pharmacogenomics J. 2013;14(16):2013–21.

33. Crespin DJ, Federspiel JJ, Biddle AK, et al. Ticagrelor versus genotype-driven antiplatelet therapy for secondary prevention after acute coronary syndrome: a cost-effectiveness analysis. Value Health. 2011;14(4):483–91.

34. Jiang M, You JH. Review of pharmacoeconomic evaluation of genotype-guided antiplatelet therapy. Expert Opin Pharmacother. 2015;16(5):771–9.

35. Khera AV, Chaffin M, Aragam KG, et al. Genome-wide polygenic scores for common diseases identify individuals with risk equivalent to monogenic mutations. Nature Genet. 2018.

36. Su J, Yu Q, Zhu H, et al. The risk of clopidogrel resistance is associated with ABCB1 polymorphisms but not promoter methylation in a Chinese Han population. PLoS One. 2017;12(3):e0174511.

37. Chen Y, Huang X, Tang Y, et al. Both PON1 Q192R and CYP2C19*2 influence platelet response to clopidogrel and ischemic events in Chinese patients undergoing percutaneous coronary intervention. Int J Clin Exp Med. 2015;8(6):9266–74.

38. Tatarunas V, Kupstyte N, Zaliunas R, et al. The impact of clinical and genetic factors on ticagrelor and clopidogrel antiplatelet therapy. Pharmacogenomics J. 2017;18(10):969–79.

39. Varenhorst C, Eriksson N, Johansson A, et al. Effect of genetic variations on ticagrelor plasma levels and clinical outcomes. Eur Heart J. 2015;36(29):1901–12.

40. Cavallari LH, Obeng AO. Genetic Determinants of P2Y12 Inhibitors and Clinical Implications. Interv Cardiol Clin. 2017;6(1):141–9.

41. Saab YB, Zeenny R, Ramadan WH. Optimizing clopidogrel dose response: a new clinical algorithm comprising CYP2C19 pharmacogenetics and drug interactions. Ther Clin Risk Manag. 2015;11:1421–7.

42. Miura G, Ariyoshi N, Sato Y, et al. Genetic and non-genetic factors responsible for antiplatelet effects of clopidogrel in Japanese patients undergoing coronary stent implantation: an algorithm to predict on-clopidogrel platelet reactivity. Thromb Res. 2014;134(4):877–83.

43. Lee JA, Lee CR, Reed BN, et al. Implementation and evaluation of a CYP2C19 genotype-guided antiplatelet therapy algorithm in high-risk coronary artery disease patients. Pharmacogenomics J. 2015;16(4):303–13.

44. Siller-Matula JM, Lang IM, Neunteufl T, et al. Interplay between genetic and clinical variables affecting platelet reactivity and cardiac adverse events in patients undergoing percutaneous coronary intervention. PLoS One. 2014;9(7):e102701.

45. Yeh RW, Secemsky EA, Kereiakes DJ, et al. Development and validation of a prediction rule for benefit and harm of dual antiplatelet therapy beyond 1 year after percutaneous coronary intervention. JAMA. 2016;315(16):1735–49.

